# Collecting health narratives at scale: A multi-study evaluation of speech-to-text in health surveys

**DOI:** 10.64898/2026.09.23.26363819

**Authors:** Andreas Baumer, Pashalis Naoumis, Jonas Fabian, Sofia Strukova, Markus Wolf, Tala Ballouz, Andrea Farnham, Valerie X. C. Hofmann, Giovanni Spitale, Volker Dellwo, Andrea Glässel, Milo Puhan, Viktor von Wyl

**Affiliations:** Epidemiology, Biostatistics and Prevention Institute, University of Zurich, Zurich, Switzerland; Digital Society Initiative, University of Zurich, Zurich, Switzerland; Department of Psychology, University of Zurich, Zurich, Switzerland; Institute of Biomedical Ethics and History of Medicine, University of Zurich, Zurich, Switzerland; Linguistic Research Infrastructure, University of Zurich, Switzerland; Institute of Public Health, Zurich University of Applied Sciences, Winterthur, Switzerland

## Abstract

Patient narratives can reveal aspects of health that standardized measures miss, yet collecting open-ended responses at population scale remains difficult. We evaluated whether speech-to-text (STT) could enable richer responses in online health surveys without the resource demands of interviews. Across four surveys involving people with post-COVID-19 condition, healthy adults, people who engage in sex work, and older adults, participants could answer open-ended questions by typing or using STT. We compared response length, lexical diversity, stop-word proportion, and total unique content words, alongside STT uptake and participant feedback. Quantitative comparisons were made using a mixed-effects model including modality (STT vs typed) as fixed effect and a random intercept per participant. STT uptake varied markedly across populations (0.7–80.0%). Spoken responses were estimated to contain more characters than typed responses in two studies, by 245.0 and 891.8 characters on average (both p<0.001), and contained more unique content words, despite lower lexical diversity and higher stop-word proportions. Participants generally valued STT for its convenience and spontaneity, but uptake was constrained by context, privacy, interface design and individual preferences. STT may enable population-scale collection of richer health narratives, provided implementation is tailored to the study population and research context.

## Introduction

Patient narratives are a cornerstone of patient-centered health research. Through free-text and spoken accounts, patients can communicate the lived realities of illness, including symptoms, functional limitations, emotional impacts, and care needs that may not be readily captured by standardized clinical measures alone (Spitale et al., 2023). Incorporating patient perspectives into research is increasingly recognized as necessary not only for patient-centered care, but also for generating valid and clinically meaningful evidence (Renjith et al., 2021).

Qualitative approaches are particularly valuable for identifying aspects of illness that lie outside existing medical frameworks (Meuleman et al., 2025). Because they allow participants to describe experiences in their own words, open-ended methods can reveal novel symptoms, unmet needs, and unexpected patterns (Ettlin et al., 2025) that structured questionnaires are unlikely to capture. This is especially pertinent in the case of chronic illnesses where symptoms wax and wane and individual experiences vary widely (O’Brien et al., 2023). Qualitative data therefore play a central role in exploratory research, hypothesis generation, the development of patient-reported outcome measures, and the contextualization of quantitative findings. However, qualitative data are expensive and time-consuming for both researchers and patients to collect, and usually only feasible for a relatively small number of participants. This has meant that incorporating qualitative patient perspectives into research at the scale of quantitative research has largely not been possible.

Post COVID-19 condition – or Long COVID –illustrates the importance of collecting diverse patient narratives concretely (Soriano et al., 2022). Early in the COVID-19 pandemic, many individuals reported persistent symptoms such as fatigue, cognitive impairment, and post-exertional symptom exacerbation, which were not well reflected in prevailing clinical descriptions of the disease. Online patient communities and qualitative accounts documented this broad range of experiences and helped shape subsequent research agendas well before these manifestations became widely recognized by researchers and healthcare systems (Au et al., 2022; Chasco et al., 2022). This example highlights a broader limitation of research that relies primarily on predefined concepts: clinically meaningful aspects of the patient experience can remain invisible until they are systematically elicited in patients’ own words.

While many health surveys include free text responses to introduce some measure of open-endedness (Singer & Couper, 2017), these often are only able to gather low-quality data, which are not suited for qualitative analysis (LaDonna et al., 2018), and often suffer from high and non-random missingness (Goldberg et al., 2025). Restricting patients to typing out their responses also introduces access barriers for some patient groups. Patients suffering from fine-motor impairments such as patients with Parkinson’s disease (De Vleeschhauwer et al., 2021), may struggle to physically type their response. For patients experiencing conditions such as fatigue or brain fog, the prolonged effort of writing long texts itself constitutes a barrier. Lower educational background is also predictive of non-response to a free text question (Rich et al., 2013), potentially indicating a task-related barrier. All told, free-text fields in surveys are insufficient for fully capturing the patient perspective.

Previous studies have described that spoken answers, captured by participants speaking into their own digital device, may provide more and better information than written answers (Höhne et al., 2024). This has, however, rarely been extended to medical research where different patient groups have varying abilities and needs when participating in studies.

In this analysis, we describe four health studies with different study populations (Long COVID patients, sex workers, adults without acute mental health issues, and older adults) which provided respondents with the option to dictate their answers. We aim to determine first, if integrating STT into online health surveys is feasible; second, how participants respond to it; and third, quantitatively compare spoken and typed answers.

## Methods

We utilized the Talk2UZH data collection platform in four cross-sectional health research surveys; three were based in Switzerland and one included recruitment across Europe. The studies were chosen to enable comparison of diverse study populations and research questions, based on an open call for interested researchers in the launch phase of the Talk2UZH tool.

### Speech-to-Text infrastructure

Talk2UZH is a voice transcription research infrastructure project, which was integrated into the Research Management Information System (RMIS), an existing online survey platform, which provides common survey functionalities (e.g., multiple choice questions, Likert scales). The transcription from voice to text is conducted using Whisper Large V3 (Radford et al., 2022), a validated open-source STT model developed by OpenAI. Both the survey website and Whisper model are hosted on servers by the University of Zurich, Switzerland. Participants are able to respond to free text questions by typing as usual or alternatively using STT dictation. Those who use STT will see the transcribed text returned to them and can correct or edit the response if desired. Once the audio file is transcribed, it is immediately deleted to eliminate potentially identifiable biometric participant information and further enhance participant privacy. Time stamps of STT transcription and editing (if applied) are logged. The STT server is accessed via an application programming interface, with communication secured via 256-bit SSH encryption.

### Study populations

In total four studies were conducted, each sampling a distinct population and providing respondents with the option to use STT. The primary findings will be published separately by the respective PIs, while the STT-related observations are reported here. Table 1 provides an overview of key demographic characteristics of each. Participants from the constituent studies were included if they provided any non-missing answer to any of the considered free text questions.

**Table 1.** Respondent characteristics of the LWPC, MemoVoice, anSWers, and SPEAK surveys.

| *Study* | *Living with Post Covid* | *MemoVoice* | *We want your anSWers* | *SPEAK* |
| --- | --- | --- | --- | --- |
| *Description of Population* | Patients with post-Covid condition | General population | People who engage in sex work | Healthy elderly people |
| *N* | 130 | 35 | 154 | 138 |
| *% female (n)* | 74.4% (96) | 34.3% (12) | 44.8% (69)<sup>1</sup> | 71.6% (96) |
| *Age, m (SD)* | 48.4 (12.0) | 31.8 (11.2) | 33.8 (8.1) | Median 66-75<sup>2</sup> |
| *STT use, n (%)* | 70.8% (92) | 80.0% (28) | 7.1% (11) | 0.7% (1) |

Note: <sup>1</sup>For comparability defined as sex assigned at birth; <sup>2</sup>Age was assessed as a categorical variable, with 86.9% of respondents to the *SPEAK* survey being 66 years of age or older.

*Living with post COVID-19 symptoms (LWPC)* investigated patient narratives of key life events and experiences and coping strategies of people affected by Long Covid (Ballouz, n.d.). The survey aimed to capture three domains of the Long Covid experiences: 1) key events and challenges participants encountered across their health, personal, and professional lives, 2) coping strategies, support systems and resources they drew on, and 3) the advice they would offer to others affected. Participants were recruited between November 2024 and February 2025 through Long Covid related studies and patient network organizations. Participants were eligible if they were aged 18 years or older, reported symptoms related to Long Covid and/or had received a clinician-confirmed Long Covid diagnosis, and had good command of German. Responses were collected primarily in Standard German (with automatic transcription into Standard German for those using STT and speaking Swiss German). The survey was tested and iteratively refined by members of the research team prior to its launch, including checks of question wording and functionality of the speech-to-text function. The study was approved by the ethics committee of the Canton of Zurich (BASEC-Nr. 2024-01025) and electronic consent was obtained from all participants prior to study participation. A preprint of the primary findings has been published (Böhm et al., 2026).

*MemoVoice* investigated how autobiographical memories differ in their representation across sensory, emotional, and structural dimensions, comparing neutral, stress-related, and reward-related memories in healthy participants. These memories collected online will then later be compared to those of participants with post-traumatic stress disorder (PTSD) and cocaine use disorder (CUD) conducted on site as part of a clinical trial. Participants were asked to recall one reward-related, one stress-related, and one neutral memory, and to describe the specific sensory-perceptual detail they could retrieve for each. In contrast to a large-scale online survey, this design is more closely analogous to an interview study.

Participants were recruited between July to December 2025 via an online participant recruitment platform, as well as through local student groups and university mailing lists. Participants were eligible if they were between 18 and 60 years of age and sufficiently fluent in German to follow the study and provide consent. Participants were excluded if they reported current psychological treatment or a psychiatric diagnosis, except for mild or moderate substance use disorder (SUD) for nicotine; lifetime exposure to a traumatic event with persistent negative outcomes; acute intoxication with alcohol or cannabis; more than 25 lifetime instances of using cocaine, amphetamine, methamphetamine, MDMA, ketamine, psychedelic, benzodiazepine, or other psychotropic substance use, including novel psychoactive substances, non-prescribed opioids, or methylphenidate; showed signs of acute mental health issues according to the GHQ-12; or self-reported suicidal ideation in the last 12 months. The study was approved by the Cantonal Ethics Committee of Zurich as an amendment to the aforementioned other clinical trial, in which the narratives of individuals with PTSD and CUD were collected prior to a pharmacological intervention aimed at attenuating intrusive memories (BASEC-Nr. 2022-01177). Electronic informed consent was obtained from all participants prior to participation.

*Share your anSWers* (hereafter abbreviated to anSWers) is a multi-country cross-sectional survey investigating how mobility shapes access to sexual healthcare and HIV pre-exposure prophylaxis (PrEP), an HIV prevention drug, among sex workers in Europe. The study was conducted through a collaboration between the University of Zurich (UZH), the European Sex Worker Alliance (ESWA), The Love Tank (London), Checkpoint Barcelona, Checkpoint Berlin, Checkpoint Zurich, Checkpoint Milan, the Institute of Tropical Medicine Antwerp, and the Public Health Service of Amsterdam. Participants were recruited between 18 January and 1 December 2026 through partnerships with sexual health services and associated community organizations using both in-person and digital outreach (e.g., QR codes on flyers and cards distributed at clinics, QR code stickers placed on PrEP packets, links shared through WhatsApp groups and mailing lists targeted to sex workers, and partnership with community-organized events). Individuals were eligible to participate if they were aged 18 years or older and reported experience of sex work, broadly defined as any consensual exchange of sexual services for money, goods, or other benefits. The study could be filled out in English, Spanish, Dutch, French, Italian, German, Portuguese, and Romanian. Ethical approval and/or ethical exemption for an anonymous survey was sought for each of the study countries as required by local law. The anSWers study is ongoing, in the present manuscript we consider answers giving up to September 22, 2026.

*SPEAK* is an online survey, which investigates the knowledge and attitudes among older Swiss people towards ICU treatment, particularly in cases where they are unable to consent. It investigates patient understanding and expectations from a medical ethics perspective. Recruitment took place through social media advertisements, and all answers were collected in December 2024. The study could be completed in German, French, Italian and English. Full details are available on the study’s OSF repository (Spitale et al., 2024). The study was submitted to the ethics committee of the Canton of Zurich (BASEC Req-2024-00649), which determined it falls outside the scope of the Human Research Act, so no formal approval was required.

#### Text processing and statistical analyses

Free text responses were tokenized, lemmatized and stop words identified using language specific models implemented in the udpipe (Straka & Straková, 2017) library and snowball stopword lists (Porter, 2001). Stopwords were retained, as their prevalence was part of the evaluation.

We compared spoken and typed responses on answer length, percentage of stop words and the number of unique lemmatized non-stop words. We further calculated the *Moving Average Type-Token Ratio* (MATTR) with a window size of 25 words. The MATTR measures the lexical diversity of a text and ranges from 0 to 1 with higher numbers indicating higher lexical diversity. It is independent of text length and robust given narrow window sizes (Zenker & Kyle, 2021). Window size of 25 was chosen as a compromise between estimate stability and inclusion of as many answers as possible.

Participant characteristics were analyzed descriptively using percentages for categorical and means for continuous variables. Text metrics were compared between typed and spoken answers within each study using a linear model including the answer modality (typed vs. STT) as a fixed effect and a random intercept for each participant. The resulting p-values were then corrected for multiple comparisons using the Benjamini-Hochberg method. For comparability between studies, answer length and number of unique content words were expressed as percentages using the average typed answer as a reference value, i.e. 100%.

#### Qualitative analysis of participant feedback

Study participants were also asked to provide feedback on the speech-to-text function. For studies that saw significant uptake issues, we conducted an LLM-assisted descriptive coding of the reasons why STT was not used. This was done using AutoCode (Spitale, 2026) an open-source tool, which applies a researcher-defined code book to participants’ answers and proposes new categories in unclear cases. For such deductive coding tasks LLM-assisted approaches have been shown to be non-inferior to human coding (Hill et al., 2026) and particularly suitable for short texts (Bermejo et al., 2025).

## Results

### Study populations

Participant characteristics and uptake of STT differed between surveys (Table 1). For this analysis we only considered participants who provided at least one response (by STT or typing) to free text questions. *LWPC* included 130 participants of which the majority were female (74.4%) with a mean age of 48.8 (SD=12). In the final analysis of *MemoVoice*, 35 participants were included, 34.3% of which were female, with a mean age of 31.8 (SD=11.1). The 154 participants included from the *anSWers* survey were highly diverse with 22.7% (n=35) of respondents being transgender. In terms of gender identity 48.1% (n=74) identified as women, 36.7% (n=56) as men and 16.2% (n=25) as non-binary. As only one participant opted to use STT in the SPEAK survey, it is not included in further evaluations.

#### STT uptake and lexical analysis of responses

Survey participants also varied strongly in STT usage. Uptake (i.e. used STT at least once) was high in LWPC (70.8%) and MemoVoice (80.0%) and low in anSWers (7.1%) and SPEAK (0.7%) surveys.

All responses to LWPC considered here were German. In MemoVoice all participants responded in German, except for one who responded in English without using STT. Responses to anSWers were diverse in language both those written (English: 45, Italian: 30, German: 21, Spanish: 21, Portuguese: 13, Dutch: 9, French: 3) and those spoken (Spanish: 6, English: 3, German: 2).

Answer length differed between surveys. Typed answers were longest in LWPC (mean (SD) = 789.1 (634.5) characters) while spoken answers were longest in MemoVoice (mean (SD) = 1472.2 (661.7) characters). Responses to the anSWer survey were shorter in both modalities with on average 98.1 (SD=112.7) typed and 245.3 (SD=291.8) spoken characters. MATTR could not be assessed in answers below 25 tokens, which excluded from LWPC 29 (4.1%), from MemoVoice 3 (2.9%), and from anSWer 249 (77.1%) responses.

Results of the quantitative evaluation are displayed in Figure 1, with model-based comparisons reported in Table 2. Across all three studies, the estimated effect of STT on response length was positive, although differences were statistically significant only in LWPC and MemoVoice. In these studies, STT responses were estimated to contain 245.0 (95% CI 110.2–379.8) and 891.8 (95% CI 599.5–1184.1) additional characters, respectively (both p<0.001). The estimated proportion of stop words was also higher for STT responses in all three studies, with statistically significant differences in LWPC and MemoVoice. Similarly, MATTR was significantly lower for STT responses in LWPC and MemoVoice, indicating lower lexical diversity. Despite this, STT responses were estimated to contain significantly more unique non-stop words in LWPC and MemoVoice (p=0.012 and p<0.001, respectively), while no significant difference was observed in anSWer (p=0.218).

**Figure 1.**
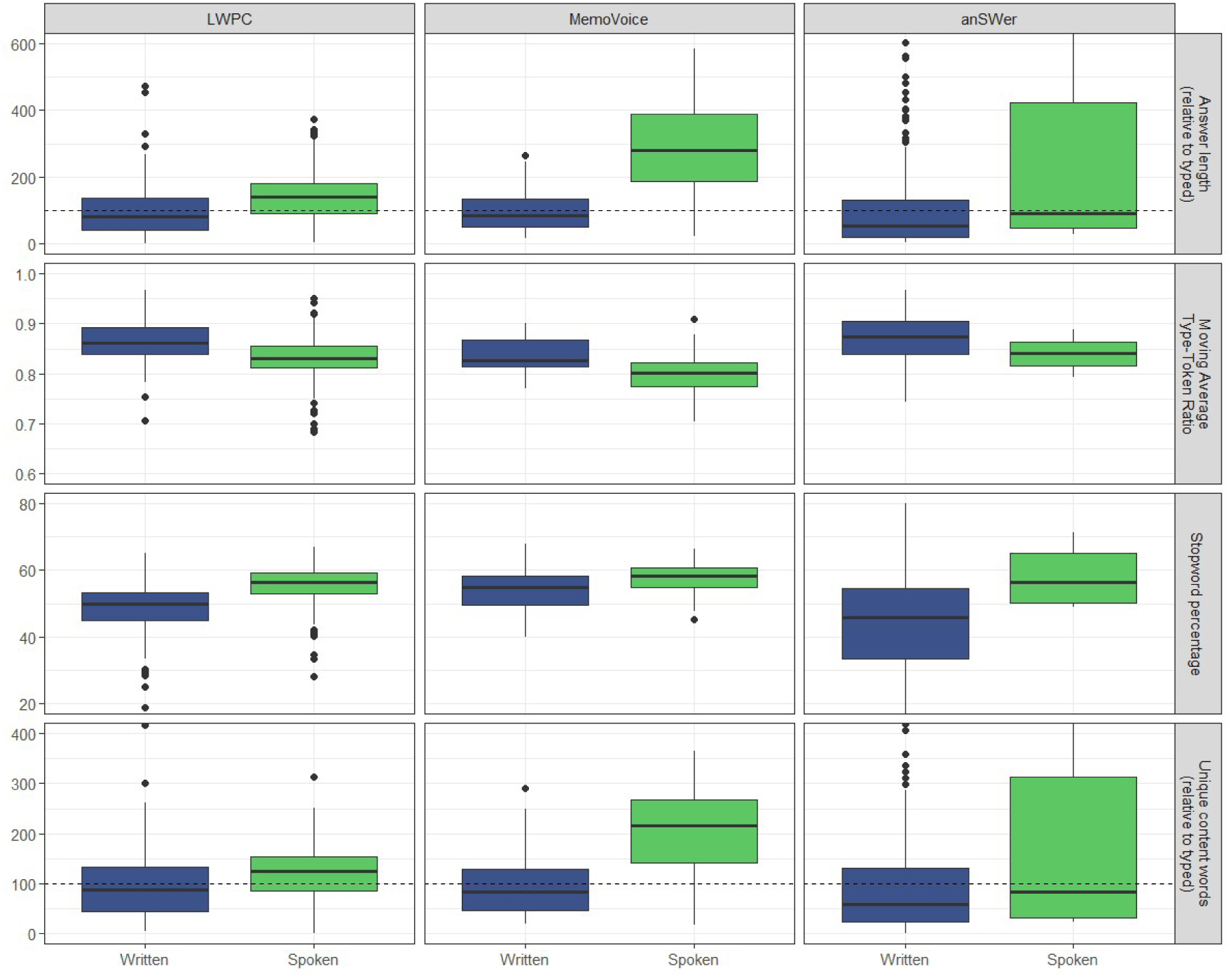
Relative answer length, Moving Average Type-Token Ratio, percentage of stop words and total unique content words compared between spoken and typed answers for the three underlying studies. Dashed horizontal lines indicate the mean value of typed answers, which was used as a reference factor.

**Figure 2.**
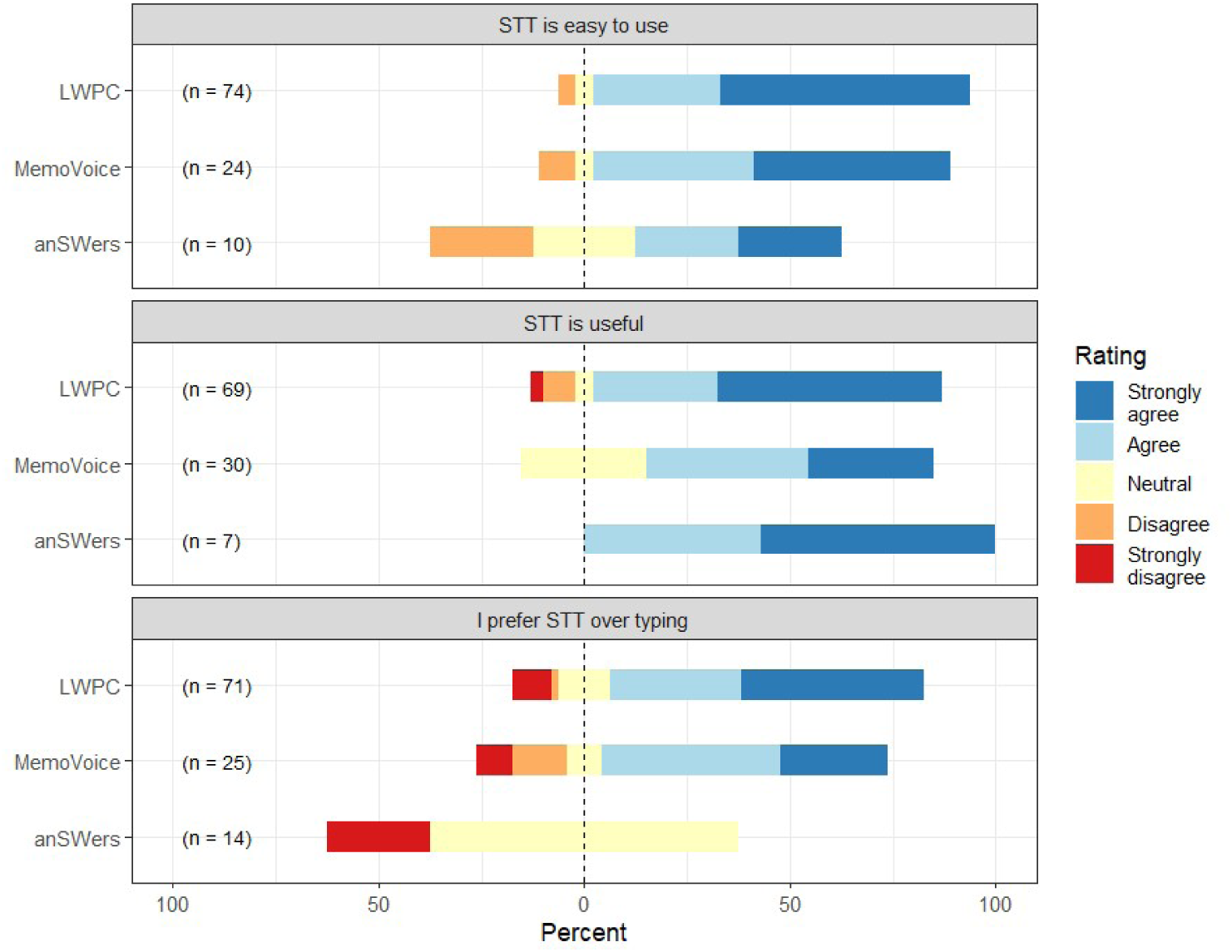
Subjective assessment of usefulness, ease of use and preference for STT across the three underlying studies.

**Table 2.** Means and p-values comparing typed and spoken answers on number of characters, Moving Average Type-Token Ratio, total unique non-stop words.

| Outcome | Survey | Typed, mean (standard deviation) | STT<sup>1</sup>, mean (standard deviation) | Mean difference[95% CI] | p-value |
| --- | --- | --- | --- | --- | --- |
| Number of characters | LWPC<sup>2</sup> | 789.12 (634.49) | 1083.80 (492.45) | 244.98 [110.19, 379.77] | <0.001* |
| Number of characters | MemoVoice | 497.16 (351.25) | 1472.24 (661.72) | 891.77 [599.47, 1184.07] | <0.001* |
| Number of characters | anSWers<sup>3</sup> | 98.13 (112.72) | 245.33 (291.80) | 95.67 [-5.13, 196.47] | 0.084 |
| Proportion stopwords in % | LWPC | 48.11 (8.54) | 55.75 (5.33) | 5.38 [3.71, 7.05] | <0.001* |
| Proportion stopwords in % | MemoVoice | 53.48 (6.15) | 57.89 (4.25) | 3.84 [1.42, 6.27] | 0.004* |
| Proportion stopwords in % | anSWers<sup>3</sup> | 41.61 (22.95) | 58.14 (9.47) | 14.68 [-4.92, 34.29] | 0.170 |
| MATTR | LWPC | 0.87 (0.04) | 0.83 (0.04) | -0.03 [-0.04, -0.02] | <0.001* |
| MATTR | MemoVoice | 0.84 (0.04) | 0.80 (0.04) | -0.03 [-0.05, -0.01] | 0.004* |
| MATTR | anSWers<sup>3</sup> | 0.87 (0.05) | 0.84 (0.07) | -0.03 [-0.11, 0.05] | 0.477 |
| Unique non-stopwords | LWPC | 52.12 (38.09) | 62.60 (25.46) | 10.22 [2.73, 17.71] | 0.012* |
| Unique non-stopwords | MemoVoice | 35.03 (25.07) | 73.85 (29.26) | 36.94 [22.88, 51.01] | <0.001* |
| Unique non-stopwords | anSWers<sup>3</sup> | 8.36 (9.10) | 16.67 (19.86) | 5 [-3.01, 13.02] | 0.240 |

Note: p-values based on a linear model including a random intercept per participant and corrected for multiple comparison using Benjamini-Hochberg method given 12 tests. <sup>1</sup>STT: Speech-to-Text; <sup>2</sup>LWPC: Living with Post-Covid; <sup>3</sup>anSWers: We want your anSWers. <sup>4</sup>MATTR: Moving average type-token ratio with window size of 25 tokens.

Overall, STT responses in all three studies included in the quantitative analysis were more extensive, less lexically diverse, had a higher stopword proportion, and more unique non-stopwords. These effects were however only significant for the two studies with high STT uptake.

#### Quantitative analysis of participant feedback

Participants who used STT generally reported positive experiences, rating it as both easy to use and useful, although perceptions varied between study populations. Participants who did not use STT were not asked about their experience with the feature. Among LWPC participants, 91.5% agreed or strongly agreed that STT was easy to use, and 84.8% agreed or strongly agreed that it was useful. Among MemoVoice participants, 86.4% agreed or strongly agreed that STT was easy to use, while 68.2% agreed or strongly agreed that it was useful.

Participants had the option to continue to edit their response after using STT, which some participants did to correct transcription errors. Across the three studies, we detected post-transcription additions (i.e. any text being entered manually after STT use) in 10.4% to 66.5% of responses (Table 3).

**Table 3.** Number of corrected and uncorrected responses split by study. Note: Editing means any type of keyboard input after using speech-to-text.

| Study | # of edited responses | # of unedited responses |
| --- | --- | --- |
| LWPC | 66.5% (n=419) | 33.5% (n=211) |
| MemoVoice | 10.4% (n=17) | 89.6% (n=146) |
| anSWer | 34.6% (n=9) | 65.4% (n=17) |

**Table 4.** Selected quotations of participant reactions to the speech-to-text option, translated and paraphrased.

| ID | Study | Quote |
| --- | --- | --- |
| Q1 | LWPC<sup>1</sup> | Saves time, less thinking, rather able to speak freely and spontaneously. |
| Q2 | LWPC | I find this speech recognition extremely helpful, because it is substantially easier than if I still had to type all this, because it really is part of the problem with the concentration, so for people with long COVID or MECFS it is extremely helpful. |
| Q3 | LWPC | I started crying and then could not talk anymore. |
| Q4 | LWPC | Typing gives me more time to think and double-check. I have always preferred it and even more so since MECFS. |
| Q5 | MemoVoice | It was faster. |
| Q6 | MemoVoice | I don't like recording myself and I think that I express myself more coherently in writing, instead of jumping back and forth. |
| Q7 | MemoVoice | I had to leave the quiet room, so I wouldn't bother anyone |
| Q8 | MemoVoice | I don't like recording myself and sending the audio to strangers. |
| Q9 | anSWer | Trans and non-binary people can be self-conscious of the sound of the sound of their voice, so it is useless. |
| Q10 | anSWer | It sucks when the microphone doesn't hear you properly and types nonsense. Especially, when it is nosy, I end up taking more time to edit the text than it would have taken to type in the first place. |
| Q11 | anSWer | I'm too lazy to type, so dictating a message, while I'm in a taxi or putting on makeup is very convenient. |
| Q12 | anSWer | It does not work for Ukrainian or Russian |
| Q13 | anSWer | Unfortunately I did not try it. Perhaps a note could have been given beforehand — before the questions start — that the voice option exists |
| Q14 | SPEAK | I hate voice-based answering! |
| Q15 | SPEAK | I don't like chatting with a computer. |
| Q16 | SPEAK | I don't like artificial intelligence. |
| Q17 | SPEAK | The interface is a bit unclear because you have a pause and stop button and it is not clear what you need to press. I took me a moment and I don't know if other participants will manage. |
| Q18 | SPEAK | I could not get it to work |

Note: <sup>1</sup>Life with Post-COVID 19 Symptoms.

The individual rate of editing texts after transcription varied greatly between respondents who used STT, with 26.6% editing every response and 40.6% editing none. The rate of post-transcription editing did not correlate with participants’ ratings of usability, usefulness or preference (r < 0.06, p > 0.46).

### Qualitative analysis of participant feedback

Participants reported a wide range of experiences and attitudes towards STT in free text responses, with differences emerging between populations. Quotations are paraphrased and translated to English if they were provided in a different language. Because participants in SPEAK and anSWers engaged with STT the least, we conducted llm-assisted descriptive coding of negative experiences and reasons why they did not use the technology. A full report on all observed codes is available in the supplementary material (sFigure 1).

Respondents to both LWPC and MemoVoice highlighted convenience (Q1, Q2, Q5) and the ability to answer more spontaneously (Q2) as major advantages. The occurrence of transcription errors and the need to double-check the transcribed text was highlighted as a disadvantage (Q9). Some participants stated that they preferred to type as it better allowed them to gather their thoughts (Q4).

LWPC respondents reported that STT was particularly useful for them as their fatigue made typing long responses taxing (Q2). However, a different respondent directly contradicted this experience, stating that STT increased their cognitive load (Q4).

STT use was very low in the SPEAK (0.7%) and anSWers (7.1%) studies. In total 103 participants of SPEAK and 82 responders of anSWers provided any negative experience or reasons why they did not use STT. Most answers did not provide a concrete negative experience or reason for not using STT (35.0% in SPEAK, 34.1% in anSWers).

In SPEAK 7.6% rejected STT out of principle with some expressing strong personal sentiments, either against speaking to a computer specifically (Q14, Q15) or artificial intelligence related topics in general (Q16). This was only the case in 1.2% responses for anSWers. Technical and effort-related content was present in both studies (11.7% in SPEAK, 8.5% in anSWers), but described different problems (Q10, Q17).

The single respondent in SPEAK who used STT successfully highlighted that it might be technically challenging for other respondents (Q17). Others replied in a manner consistent with experiencing technical difficulties (Q18).

Because the anSWers survey primarily recruited participants in community spaces many people filled out the survey there and reported problems with being overheard or being embarrassed to speak out loud. One participant also responded that they disliked STT and believed it to be an unsuitable modality for gender minority people because their voice does not match their desired gender expression (Q9).

Responses referring to non-use and non-adoption were frequent in anSWers as in SPEAK (46.3%). Thirty-four anSWers respondents (41.5%) stated that they had not used the function without offering any reason (Q12) is typical of the whole group. One participant also noted that they did not realize that STT was an option (Q13).

## Discussion

Our multi-study evaluation suggests that voice-based response entry can substantially expand the amount of text generated in online health surveys. By contrasting the uptake and response of participants to STT between studies, we provide initial insights into the design and context factors which influence its feasibility and utility. Among participants who chose STT, responses were substantially longer and despite lower lexical density and higher stop word ratios, they contained more unique non-stop words overall. This pattern suggests that speaking may encourage more in-depth responses. We further find evidence that STT can provide improved accessibility to certain participant groups, such as those suffering from fatigue.

This finding is particularly relevant to population-scale health research, where conventional surveys have to find a compromise between capturing the complexity of participants lived experience and scalability. By reducing the effort required to produce extended free-text responses, voice-based response entry may allow population-scale digital studies to better integrate patients’ lived experience.

Out of four studies we characterize the inclusion of STT as successful in two cases (LWPC, MemoVoice), partially successful in one case (anSWers), and unsuccessful in one case (SPEAK), despite the technology in question being largely identical. We will first discuss the implications for STT in health research for each study individually and then elaborate on the more general insights, limitations, and implications.

*Living with Post Covid-19 symptoms* participants responded most enthusiastically to STT as survey modality, with a majority opting to use it and rating it as useful and easy to use. This study population is characterized by an ongoing effort to have their condition understood and recognized by the medical community and was highly motivated to make their voice heard. Their high motivation was also made apparent by the long answers provided by those who chose to type and the high rate of edits after using STT. Interestingly, some participants stated that STT was especially useful because fatigue makes typing a chore, while others reported the opposite (i.e. STT not being a suitable option due to fatigue). This highlights the heterogeneity in responder preferences when it comes to questionnaire modality even within the same population.

*MemoVoice* proved a strong natural fit of the research question with STT-based surveying because participants were tasked with recounting multiple recent emotionally charged memories at considerable length. Thus, most opted to use STT to provide these narratives, which increased the length of responses, as well as self-reported convenience and spontaneity. In the absence of STT, this study would have been conducted as an interview study, which would have greatly increased the resource demand for the research team and introduced known accessibility barriers (Harris & Roberts, 2003).

*We want your anSWers* highlighted the potential for research access and scalability but also the critical importance of contextual factors. Because recruitment took place at in-person locations for sex workers such as night cafes, participants were not in a space where they felt comfortable speaking, especially about the highly sensitive topics at the core of this research. The space where patients respond to such research needs to be considered in study design. As the study continues to collect data, some recruitment localities will attempt to provide such spaces. Despite these challenges, it is a promising sign that some in-depth responses could be gathered from sex workers at risk of HIV exposure, who are a hard-to-reach and highly multilingual research population (Barros et al., 2015).

Survey modality and its effect on sex workers’ likelihood to disclose high risk behaviors have been previously explored in the form of audio computer assisted self-interview (Brown et al., 2013). While in these approaches respondents still type their answers, it is a useful point of reference as it has been shown that sex workers are more likely to disclose important but sensitive details, when they are able to self-interview comparing to responding to an interviewer (Phoo et al., 2022).

*SPEAK* respondents, an older general-population sample, largely declined to use STT and those who tried it encountered technical challenges, in part due to a non-sample appropriate introduction and design of the interface. Some even articulated that they fundamentally dislike speaking to a machine. It should be noted that the written responses still supported a full thematic analysis, meaning that these challenges are not due to low motivation overall. This underscores how important it is to consider user design aspects, when introducing new modalities into research, especially when the target population is not already familiar with or open to the new feature. It would be particularly useful if words appeared as people spoke, instead of the full answer needing to be recorded and transcribed in one.

In summary, we identify the following factors as important enablers to STT in health research. First, the study and question design should be oriented around capturing patient narratives rather than checking symptom lists. Second, STT needs to be introduced in a manner that is clearly understandable and adequate for the study population. Third, participants should be in a calm and private environment when responding to the survey, so they feel comfortable speaking out loud. Fourth, members of the study population should be sufficiently motivated to engage with a novel form of data collection and optimally be inclined towards STT or similar technologies, such as voice notes.

Our finding that spoken responses were more extensive and contained more unique content words despite lower lexical diversity aligns closely with previous research. This research was however conducted in political science and had not previously been extended to health research (Gavras et al., 2022; Höhne et al., 2024). In contrast to our design, these studies randomized respondents to either speaking or typing conditions, which increases the comparability of answers as it eliminates self-selection biases (e.g., more motivated people may be more likely to type). The fact that our findings align with research absent gives us some confidence in the internal validity of our analyses, while being closer to how STT might be implemented in practice.

It is important to be conscious of potential emerging digital divides, and to enable all patient populations to equally benefit from the advantages of modern STT capabilities. Access to a microphone of adequate quality and a stable internet connection are required, meaning that patients with low-cost or older devices may receive more error-prone transcriptions than those with higher quality devices. Further, STT explanations should be tailored for patients with lower digital literacy. Lastly, while many primarily digital surveys provide pen and paper alternatives if desired, an equivalent alternative STT (i.e. dictation in person) may not be feasible at scale.

While in our studies only one participant reported rejecting STT due to gender dysphoria-related dislike of their voice (Q9), it highlights that STT is only a beneficial modality to participants who are comfortable and able to speak. Automatic speech recognition technology is long known to constitute an accessibility barrier to people who are hard of hearing or deaf (Glasser et al., 2017). As researchers it is thus important to explicitly solicit the viewpoints and narratives from those who do not benefit from STT.

Despite these words of caution, we see the introduction of STT as an avenue to reduce existing barriers to patients making their voice heard in health research. Health research in its current form tends to overrepresent patients with high socio-economic status and education as well as men and racial majority groups (Khan et al., 2020). In many trials, fluency in the local language is explicitly required to participate (Muthukumar et al., 2021). Multilingual STT has the potential to lower the hurdles for patients who do not share a language with the research team. Further, patients who do not routinely use a keyboard (for example, as part of their job) may feel more comfortable contributing their perspective if STT is made available to them.

### Limitations and Strengths

Our choice to delete the recordings after transcription made it impossible to independently verify the accuracy of the STT model or to make retrospective corrections to responses. While this is a drawback, we believe that it helped increase participant trust.

Quantitative assessment of lexical diversity was challenging, as most established metrics require longer texts to provide stable estimates and many are dependent on text length, which is also confounded by choice of modality (speaking or typing). Many answers were too short for reliable estimation of lexical diversity and MATTR with a window size of 25 tokens was chosen as a compromise to not exclude too many. While this metric is independent of answer length, the window size is narrow, which may lead to unstable estimates.

While both LWPC and MemoVoice were mostly completed in German, anSWers contained a variety of different languages. Because lexical measures can differ systematically between languages, comparison of these metrics between studies must consider the language composition.

This was not a randomized evaluation of response modality, and participants self-selected whether to respond by typing or using STT. Differences between typed and spoken responses therefore cannot be interpreted causally and may partly reflect characteristics of the participants or questions for which STT was chosen.

A clear strength of this analysis is the inclusion of four separate studies, with varying sample populations, which allowed for observational comparisons of participant responses. However, because the four evaluation studies differed in several regards at once, our ability to identify which contextual factors were responsible for differences in STT uptake, is limited. Further, it is possible that the recruited participants are not representative of the underlying population in regard to digital literacy or other relevant characteristics.

### Future directions

Future research is needed to systematically evaluate how STT can improve the collection of unstructured health data and how this change in modality impacts the produced texts. Further, as we have shown, there are great variations between and within study populations in their adoption of STT. It is therefore important to continually evaluate under what circumstances STT can produce benefits and how it can be made accessible for study populations who may be unfamiliar with voice-based self-recording. Given the importance of design and appropriateness to the target population, we see significant potential for co-design approaches in this realm.

## Conclusion

In our four-study evaluation, health survey participants who used speech-to-text (STT) provided longer responses and, in two studies, significantly more unique content words, despite higher stop-word proportions and lower lexical diversity. At the same time, the large variation in uptake across studies demonstrates the importance of implementation context. Successful use of STT requires research questions that invite narrative responses, participants who are willing and able to speak, an environment in which they feel comfortable doing so, and interface design tailored to the study population. We see considerable potential for STT to help bring participants’ voices into population-scale health research, particularly where typing may constrain the ease or extent of open-ended responses. The multilingual capabilities of modern STT models may further broaden participation by reducing language-related barriers to open-ended data collection. By reducing the effort required to provide extended responses, voice-based data collection may enable digital health studies to capture participants’ lived experiences in greater depth without sacrificing scalability.

## Data availability statement

The text evaluation metrics for individual answers will be made publicly available at time of submission and are available upon reasonable request. The raw text data cannot be made publicly available due to privacy concerns and the sensitive nature of the data.

## Code availability statement

The code used to process the text data is available upon request, though some data manipulation scripts are excluded as they would reveal parts of the underlying data.

## Acknowledgements

The MemoVoice study was conducted within the framework of a larger project, in which Amelie Zacher and Lina Dietiker were centrally involved. We thank them for their contributions.

